# Implementation of an Enhanced Medication Access Workflow within a Health System Specialty Pharmacy: Impact on Patient & Clinician Experience

**DOI:** 10.1101/2024.05.28.24307823

**Authors:** Brandon Harkonen, Anthony Cuttitta, James Henderson, Valerie Gavrila, Jey Mckibbin, Sherrie Luttman, Wendy Benedict, Lindsey R. Kelley, Scott A. Flanders, Hae Mi Choe, Geoffrey D Barnes

**Author notes:** **Corresponding Author:** Anthony Cuttitta, MPH, 777 E. Eisenhower Parkway, Suite 600, Ann Arbor, MI 48108-3258. **Author contributions:** HMC, BH, LK, JM, SL, SF contributed to intervention design concept and implementation. BH, AC, JH, GB wrote the manuscript. GB, JH, AC, VG contributed to analytic and evaluation approaches. VG, WB, JH, and AC contributed to data acquisition & collection. JH and GB contributed to statistical expertise. All authors contributed by providing critical revision of the manuscript.

## Abstract

**Purpose:** The initiation of specialty medications is associated with patient access challenges and clinician burden. This intervention invested resources upstream of the prescription being written. The evaluation assessed the impact on patient and clinician experience.

**Methods:** The specialty pharmacy intervention was designed to improve medication access within five medical specialty clinics by utilizing an embedded medication access team assigned to patients and prescribers of targeted medications. We used a multi-methods evaluative approach. Semi-structured qualitative interviews provided an understanding of clinician experience. To quantitatively assess associations between the referral process and patient experience, we analyzed the emotional valence of patient portal messages using a retrospective cohort study and an event study framework of a non-randomized, stepped wedge implementation design.

**Results:** The intervention was associated with an increase in the net positive emotional valence of patient portal messages (AME, 5.3; 95% CI, 3.8-6.8). Except for gastroenterologists seeing patients for irritable bowel disease, patients cared for in all other specialties experienced statistically significant increases in net positive valence in the primary analysis. Regarding clinician experience, four major interrelated themes emerged from 17 qualitative interviews with prescribers and pharmacists: (1) decreased clinician burden, general praise, (2) improved experience & satisfaction, reduced anxiety & concerns, (3) rewarding praise for other prescribers/colleagues, and (4) excellent coordination, efficiency, and speed.

**Conclusion:** Investing staff resources before, during, and after the prior authorization process greatly improves clinician experience. The positive valence of patient portal messages also increased suggesting patient experience improvements.

## Introduction

Specialty medications can offer life-changing outcomes for patients. However, cost and the complexity of initiation often create challenges for patients and clinicians. Costs for specialty medications are high, acting as a barrier to access and adherence which drives specialty drug abandonment.^i,ii^ This is associated with worse patient outcomes and quality of life.^iii,iv^

In addition to patient-related specialty medication access difficulties, clinical staff struggle with the administrative challenge of prior authorizations (PAs) required by health plans to qualify for medication payment. PAs represent a primary source of dissatisfaction for prescribers and clinical staff.^v^ PAs create challenges for prescribers and clinic staff. In a recent survey by the American Medical Association, 88% of prescribers describe the burden attributed to PA as high or extremely high, with practices requiring 2 business day of staff time to complete 45 PAs per week on average.^vi^ In another survey of gastroenterologists, 93.8% of respondents perceived a high burden of PA requests, and 57.8% avoided talking about a preferred medication with their patients because of a high perceived likelihood of coverage denial. ^vii^ Extra time spent on these PAs by medical staff is also not even associated with PA approval.^viii^ Others have suggested that further research is needed to decrease the time and burden associated with PAs. ^ix^

There has been an abundance of interventions and research addressing the clinical impact of PA requirements, as well as the regular use of medical assistants, nurses, and pharmacists to address PAs.^x,xi,xii,xiii,xiv,xv,xvi^ However, these interventions rarely invest resources upstream of the prescription being written for a medication likely to require PA. In this paper, we describe a quality improvement intervention at our institution that invested resources prior to a prescription for a specialty pharmacy medication being sent to the pharmacy. We focus on evaluating the care pathway’s impact on patient and prescriber experiences.

## Methods

### The Intervention

This specialty pharmacy intervention focused on improving the medication access process for patients and prescribers within five medical specialty clinics at an academic medical center. The primary intervention included the use of an embedded medication access team assigned to patients and their prescribers of targeted medications within specialty clinics. Adjacent clinical pharmacists provided clinical specialty pharmacy support to the medication access and clinical teams. A key innovation of this care pathway is the involvement of these roles throughout the PA process, particularly before a prescription is written and sent to the pharmacy (Figure 1).

**Figure 1.**
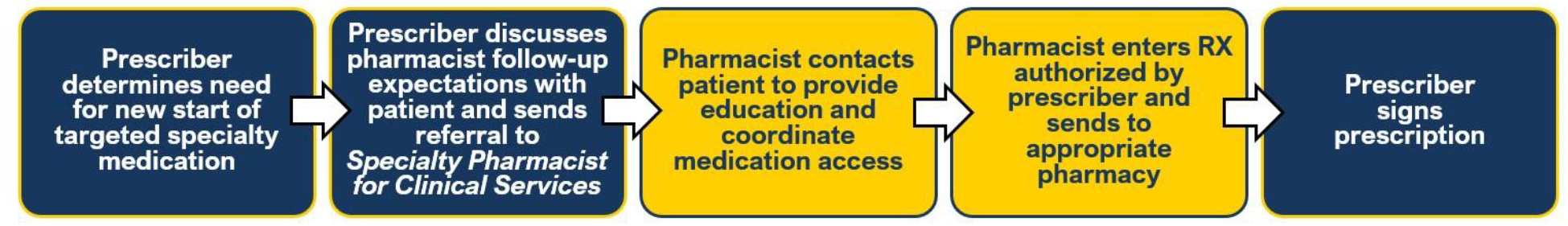
Intervention process for medication access. Multiple roles were involved throughout the prior authorization and medication access process. The pharmacist is involved prior to prescription completion.

Specifically, when the prescriber identifies the need for initiation of a qualifying specialty medication, they send a referral to a clinical specialty pharmacist in lieu of a prescription. Within the referral, the prescriber indicates the preferred medication name, dose, and frequency. After receiving the referral order, the pharmacist contacts the patient to provide telephonic clinical education and coordinate medication access in partnership with medication access staff. The medication access team completes benefit investigation, submits prior authorization, coordinates financial assistance, and identifies and documents the patient’s preferred or required network pharmacy within the medical record. The pharmacist then enters a prescription order for the specialty medication and pends the order for prescriber signature. In some cases, the medication needs to be changed following benefit investigation, which is also recommended by the pharmacist. Once resolved, the prescriber then reviews and signs the prescription order, releasing it to the appropriate pharmacy for fulfillment.

### Study Analytic Design

We used a multi-methods evaluative approach, including qualitative evaluation of the provider experience and quantitative assessment of changes in patient experience associated with the specialty pharmacy referral intervention.

#### Prescriber and Pharmacist Qualitative Evaluation

For the clinician experience, we conducted individual semi-structured virtual interviews with both physician prescribers and pharmacists across all targeted specialty services participating in the intervention. Interviews focused on PA, clinician effort, prescriber-to-pharmacist communication, and overall clinician experience. Interview content was developed from key informant discussions from numerous stakeholders familiar with the PA process, prescribing providers, and specialty pharmacy.

Recruitment of participants was accomplished through purposive sampling within two key stakeholder groups, prescribing physicians and pharmacists directly involved in the intervention program between March 2023 and May 2023. For prescribers, participants were sampled based on two variables. First, we included at least one participant from each participating clinic, including gastroenterology, neurology, rheumatology, pulmonology, and dermatology. Second, we sampled across low-use, medium-use, and high-use prescribers. For pharmacists, convenience sampling was used to interview pharmacists in different clinics. Participants were invited to participate by email, and it was made clear that participation was entirely voluntary. Additional details about the semi-structured interviews and analysis are provided in the supplemental appendix.

#### Patient Experience Evaluation

To assess associations of the referral process with patient experience, we analyzed the emotional valence of patient portal messages using a retrospective cohort study of a non-randomized, stepped wedge implementation design. ^xvii,xviii,xix^ We identified a cohort of patients between January 2, 2020, and March 28, 2023, who were either (a) first prescribed a medication for which a referral to specialty pharmacy was encouraged or (b) referred to specialty pharmacy. In both instances, we excluded patients who had previously been prescribed a study medication within our health system. We identified all portal messages sent by these patients within 90 days of the initial referral or prescription. The primary outcome was the emotional valence of these messages scored as described in the supplemental appendix. The unit of analysis was the message with the event-study (rolling pre-post) exposure defined at the provider level with patients attributed to the referring or prescribing provider.

We summarized continuous variables using means and inter-quartile ranges and categorical variables using percentages. Descriptive comparisons were made using 95% confidence intervals (CIs) from t-tests or differences in proportions. Specialty adjusted comparisons and associations of patient characteristics with exposure level were made using multivariate linear or logistic regression.

We used a linear mixed model to compare the net positive valence between the pre and post periods, defined at the provider level, while controlling for specialty and patient-level differences. Specifically, the independent variables in this model were a binary indicator for the referring or prescribing provider being in the post period, binary indicators for specialty (reference = Dermatology), and interactions between indicators for the post-period each specialty. The model also included a Gaussian random intercept for patient to account for dependence between messages from the same patient. We estimated model parameters using maximum likelihood as implemented in the lme4 package.^xx^ We used average marginal effects (AMEs) to summarize differences between pre-and-post. The AME is the average difference in the predicted net positive valence of each message between a prediction made by assuming the message occurred pre intervention and the prediction made assuming it occurred post intervention.^xxi^ For each specific message, one of the two predictions is counterfactual.

We used a similar approach using mixed logistic regression to model the change in the percentage of messages classified as positive, which we defined as those having a net positive valence above 75. The threshold was chosen by examining histograms, with most messages clustering above 90, below -90 or between -25 and 25.

In a sensitivity analysis, we assessed each specialty for pre implementation trends in the outcome using models that separately included either a common or different pre and post trends by specialty, keeping trends that significantly improved model fit as measured by the Akaike Information Criterion and a likelihood ratio test. Finally, to assess whether our results may have been confounded by unrelated differences in emotional valence during the COVID-19 pandemic, we conducted an additional sensitivity analysis excluding all messages sent during 2020.

This project was deemed exempt by the University of Michigan Institutional Review Board as a quality improvement project.

## Results

### Patient Population Impacted by Intervention

Between January 2, 2020, and March 28, 2023, 6,888 adult patients were first prescribed one of the specialty pharmacy medications or referred to specialty pharmacy. Overall, these patients had an average (IQR) age of 47.9 (35.3-59.5), 4,838 (70,2%) were female, 228 (3.3%) were Hispanic, 6,495 (94.3%) non-Hispanic, and 165 (2.4%) of Unknown ethnicity. These patients self-reported their race as follows: 41 (0.6%) American Indian or Alaska Native, 158 (2.3) Asian (includes Asian, Asian Indian, Chinese, Korean, Filipino and Other Asian.), 574 (8.3%) as Black, 5,685 (82.5%) as White, 342 (5.0%) as Other (includes Middle Eastern/North African, Native Hawaiian, Native Hawaiian or Other Pacific Islander, Multi, and Other), and 88 (1.3%) as Unknown.

From May 7, 2021, to June 7, 2022, prescribing providers in five specialties, consisting of Dermatology, Gastroenterology (Irritable Bowel Disease), Neurology (Headache and Multiple Sclerosis), Pulmonology, and Rheumatology, were invited to begin referring patients to specialty pharmacy using a non-randomized, step-wedged design (Figure 2). In total, the evaluation encompassed 335 providers, including 258 pre intervention and 242 post intervention.

**Figure 2.**
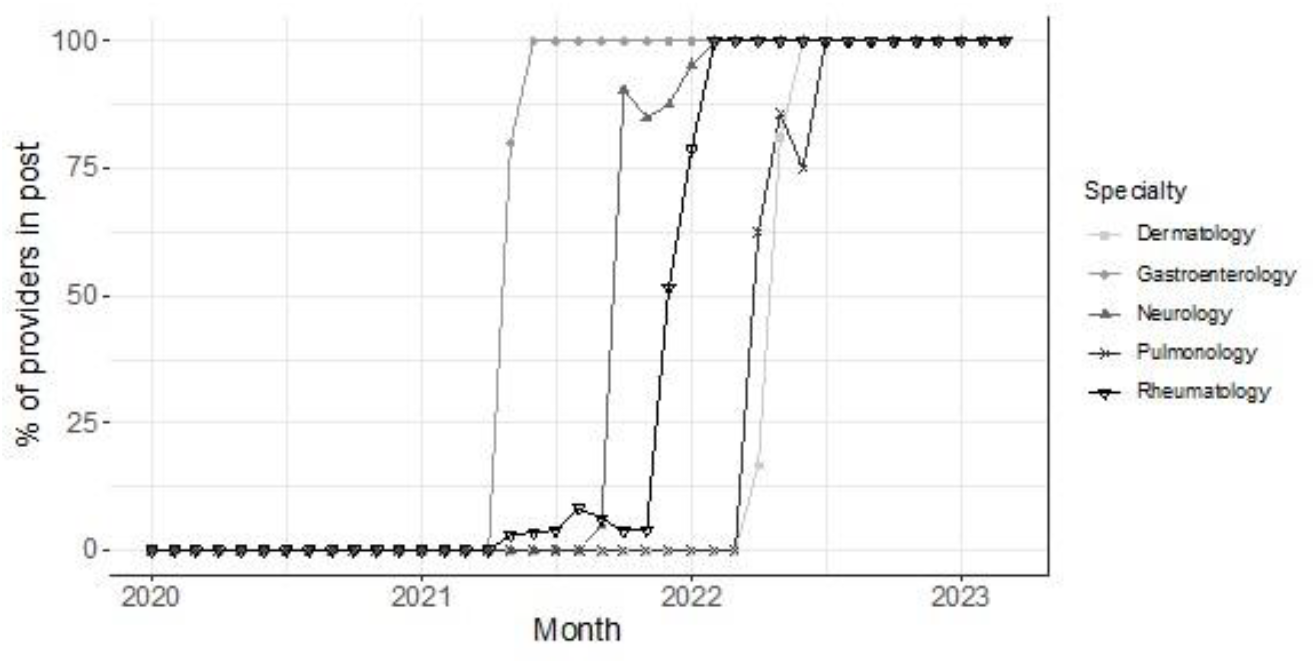
Monthly percent of referring or prescribing providers in the post period by specialty. Small, temporary declines observed for some specialties represent months in which a provider in the post period did not make a referral or an initial prescription for an in-scope specialty pharmacy drug (eTable 8).

### Patient Experience

In total 62,270 portal messages were sent by these patients in the 90-day period after the referral process or prescription began. This included 31,290 (50.2%) messages from 2,415 patients in their provider’s pre-intervention period and 30,980 (49.8%) messages from 2,131 patients in their post-intervention period. Because of the timing of how the new referral process was implemented across the various specialties (Figure 2) patients sending portal messages in the post period were more likely to be attributed to Gastroenterologists seeing patients for irritable bowel disease or Neurologists specializing in headache or multiple sclerosis. After adjusting for specialty, the post-intervention period had 2.3 percentage point (pp) (95% CI, 0.2-4.4 pp) fewer White patients, 1.0 pp (95% CI, 0.5-1.9 pp) more Asian patients and 1.5 pp (95% CI, 0.3-2.8 pp) more patients with a race designation aggregated into Other. See Table 1.

**Table 1.**
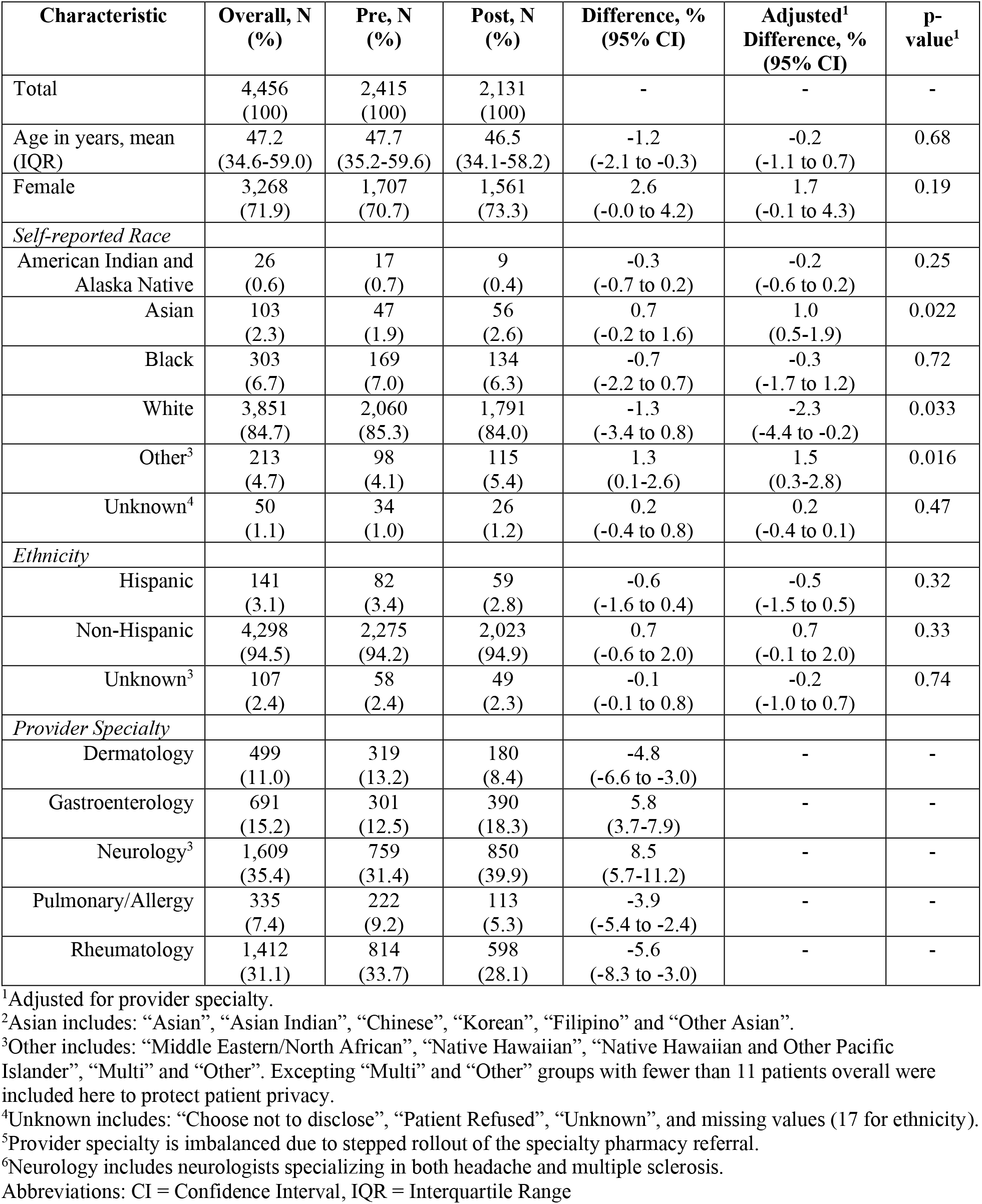
Demographics and Clinical Characteristics of Patients with Portal Messages.

After adjusting for specialty, patients in the post period were more likely to report their race as Asian or as a category aggregated into Other and less likely to report White (eTable 1). Among 2,945 patients represented in the post period, providers used the specialty pharmacy referral for 1,891 (64.2%). Referred patients were more likely to be Asian or of other race and less likely to be White than patients prescribed a specialty medication without a referral (eTable 2).

**Table 2.** Prescriber and Pharmacist Interview Participant Characteristics.

| Clinic | Clinician Type |  | Opportunities with Target Rx | Number of Referrals | % Referral Use |
| --- | --- | --- | --- | --- | --- |
|  | Prescriber | Pharmacist |  |  |  |
| Dermatology | 2 | 1 | 23 | 17 | 74% |
| Gastroenterology | 2 | 1 | 123 | 72 | 59% |
| Neurology | 2 | 2 | 336 | 255 | 76% |
| Pulmonary/Allergy | 3 | 1 | 46 | 41 | 89% |
| Rheumatology | 1 | 2 | 24 | 22 | 92% |
| **Total** | **10** | **7** | **552** | **407** | **74%** |

Using a pre-trained machine learning algorithm, we assessed the net positive valence of these messages which ranges from -100 (most negative) to 100 (most positive) with 0 representing an emotionally neutral valence. The unadjusted mean (IQR) net positive valence was 7.8 (0.0-25.0) in the pre-intervention period and 12.2 (0.0-33.3) in the post-intervention period (eTable 3). After controlling for provider specialty and patient, the intervention was associated with an increase in the net positive emotional valence of patient portal messages (AME, 5.3; 95% CI, 3.8-6.8; Figure 3 and eTable 4). Except for gastroenterologists seeing patients for irritable bowel disease, patients cared for in all other specialties experienced a statistically significant increases in net positive valence in the primary analysis.

**Table 3.** Prescriber and Pharmacist Quotes.

| Theme | Exemplar Quote |
| --- | --- |
| Decreased clinician burden, General praise | *"I think it saves both providers and patients so much time. Like I prescribe these medications very frequently, like multiple times a day. So, if I had to be a lot more involved in the prior auth process, like writing a note to a pool of auth people like these are all the medicines... It would take it at least an additional hour a day or every other day to do that work."* (Neurology Prescriber)<br>*"Yeah, I mean, it is like 1,000 times better now, I mean, it is amazing. It's like, probably like the thing that has made my life the easy like easiest out of like all the things."* (Dermatology Prescriber)<br>*"I think it just helps with it's just one less thing for providers to worry about, one less thing for patients to worry about like they don't have to keep track of it like we have a team that will look into it."* (Rheumatology Pharmacist)<br>*"The providers appreciate it because it's less it calls to them, if it's side effects and things like that, I mean, we can help sometimes manage those things potentially."* (Pulmonology Pharmacist) |
| Improved experience and satisfaction, reduced anxiety and concerns | *"The there's definitely more status job satisfaction having these relationships with the clinics and the providers."* (Pulmonology Pharmacist)<br>*"That feels good, too, to be able to do that, and them just not even have to worry about it. No, we're taking care of it."* (Pulmonology Pharmacist)<br>*"I know that I'm sending it now to specialty pharmacy, I have like no concerns whatsoever that it's going to fall through the cracks."* (Pulmonology Physician) |
| Rewarding praise for other providers and colleagues | *"I mean, you know, pharmacists have a whole, whole upper level of training...And so you know it's been very, very helpful to have you know, some someone at their level to help us with all this, you know it's been incredible."* (Neurology Prescriber)<br>*"We always are like, just, you know, throwing confetti in the air when we talk about the specialty pharmacy."* (Dermatology Prescriber) |
| Excellent coordination, efficiency, and speed | *"Just being able to feel like 'hey, like this process is efficient,' like, you know. The patient talked to the to the provider like yesterday, and within like a week they may have the drug in their hand, which previously took so long for them to get it."* (Rheumatology Pharmacist)<br>*"And the turnaround time has been incredible. I mean, you know, we've probably cut our prior authorization time down from, you know 10 to 20 days to maybe just, you know, a few days in some instances."* (Neurology Prescriber) |

**Figure 3.**
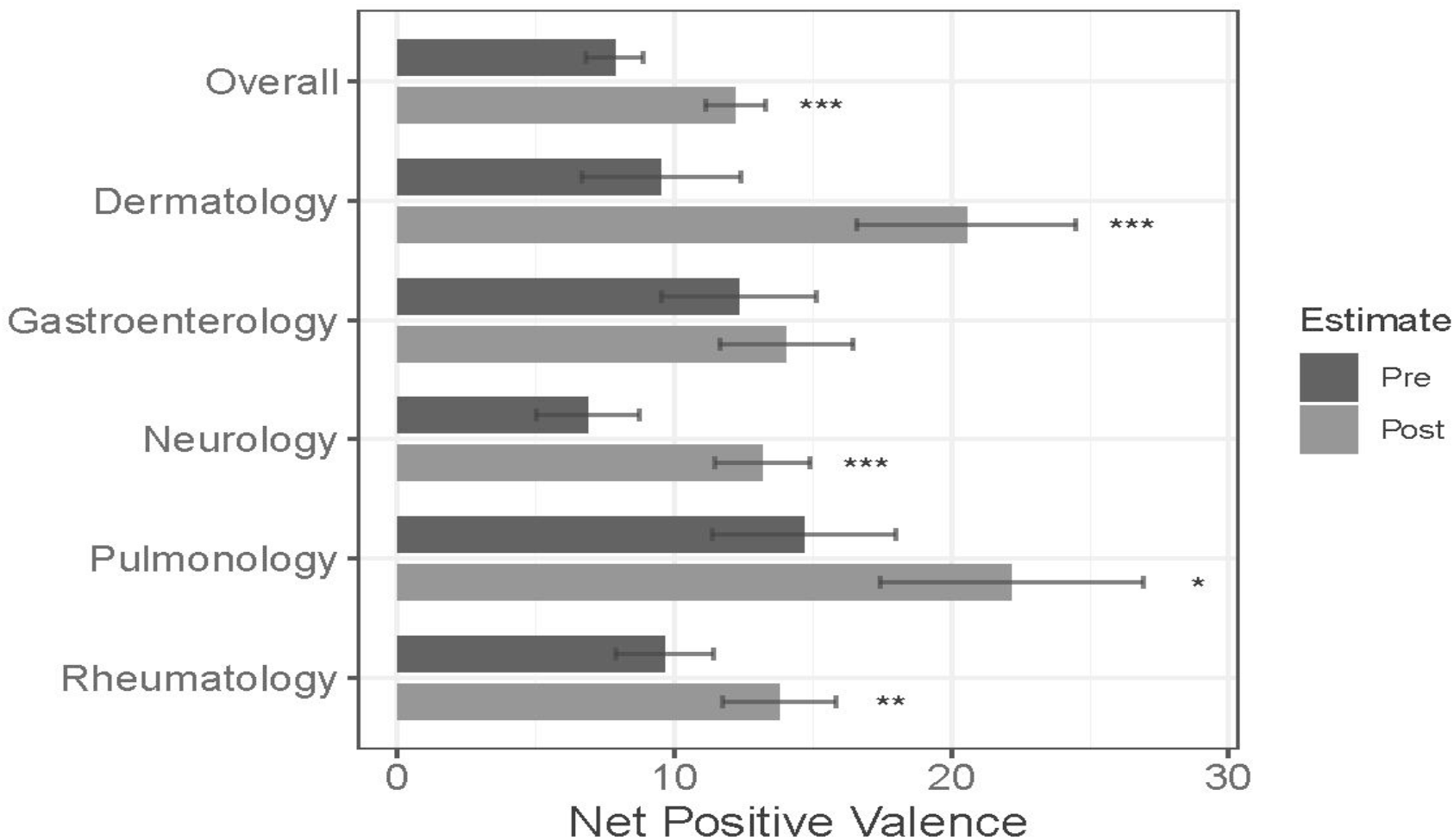
Average net positive emotional valence of patient portal messages before and after implementation. Stars indicate statistically significant differences between pre and post in the primary model at levels * < 0.05, ** < 0.01, *** < 0.001.

In a sensitivity analysis, we controlled for pre-implementation trends, the overall improvement was similar (AME, 5.1; 95% CI, 3.3-7.1) but the increase for Dermatology was no longer statistically different from zero (eTables 4 and 5). In an additional sensitivity analysis excluding all message from 2020 (due to the COVID-19 pandemic), the overall effect was smaller (AME, 3.9; 95% CI, 1.9-5.8) but only Neurology and Rheumatology had statistically significant estimated increases.

To help with interpretation, we classified messages as “positive” (75-100), “neutral” (−75 to 75) or “negative” (−100 to -75) using the net positive valence. From pre- to post-implementation, the percent of positive messages increased from 5.5% to 11.9%, neutral messages decreased from 88.4% to 80.8%, and negative messages increased from 6.1% to 7.2%. In a multi-level logistic regression comparing the percent of messages with positive valence vs negative or neutral valence and controlling for pre-implementation trends, the estimated increase in the percent of positive messages was 6.6 percentage points (95% CI, 6.1-7.7 pp). In sensitivity analyses excluding data from 2020, the estimated increase was 5.9 percentage points (95% CI, 5.0-6.9 pp). All specialties, except Dermatology, had significant increases in the percent of positive messages in analyses including and excluding 2020 (eTable 7).

### Prescriber Experience

Ten prescribers and seven pharmacists involved in this specialty pharmacy intervention consented to semi-structured interviews. Prescribers participated in 552 total opportunities with target drug prescriptions. Additional participant characteristics for both prescribers and pharmacists are described in Table 2.

#### Major Themes, Subthemes, and Codebook Development

Inductive coding led to the finalization of 24 codes defined in a codebook and finalized prior to interview coding. Once coded, key themes and excerpts were identified for the most relevant topics.

Four major interrelated themes about the prior authorization (PA), the medication access process, and clinician’s experiences emerged from interviews: (1) decreased clinician burden, general praise, (2) improved experience & satisfaction, reduced anxiety & concerns, (3) rewarding praise for other prescribers/colleagues, and (4) excellent coordination, efficiency, and speed. These themes all describe how the investment of resources early in the PA process, particularly before sending a prescription to the pharmacy, is unique and can positively impact the experience of clinicians. Exemplar quotes are included in Table 3.

##### Theme 1: Decreased clinician burden, general praise

Both prescriber and pharmacist interviewees consistently described that this specialty pharmacy intervention reduced their burden and saved a significant amount of prescriber time. In particular, they described that this process successfully shifted medication access work from the prescriber and their clinic to the medication access team for specialty pharmacy resulting in numerous benefits.

For prescribers, the intervention resulted in a clear reduction in the amount of time and administrative burden tied to medication access. Specialty clinical pharmacists also described reduced medication access and PA issues along with less burden on prescribers. They also recognized an important role in assist patients with any issues that arise specialty drugs that reduce the time and burden previously experienced by prescribers.

##### Theme 2: Improved experience & satisfaction, reduced anxiety & concerns

This intervention maximized pharmacist expertise as a member of the care team. Specifically, pharmacists valued integration early in the PA and medication access process and the subsequent improvement in clinician experience and satisfaction. This directly leads to high pharmacist job satisfaction and a recognition by prescribers that they could rely on the pharmacists without having to be concerned with the PA process and medication access for their patients.

##### Theme 3: Rewarding praise for other providers and colleagues

Prescribing physicians shared overwhelmingly positive views about the involvement of the pharmacists. Specifically, they acknowledged the direct positive impact of the pharmacists’ efforts on both prescribers and patients.

##### Theme 4: Excellent coordination, efficiency, and speed

Interviewees identified early involvement of pharmacists and financial coordinators (before a prescription is submitted) as a key driver of a positive prescriber experience. They also noted the early involvement of these team members significantly helped patient improve the speed of prescription fulfillment and provide medication access support.

## Discussion

This evaluation of an enhanced medication access workflow implementation demonstrates how health systems and specialty pharmacies can strategically utilize resources to improve the prior authorization (PA) process in a manner that positively impacts both patient and clinician experience.

Analysis of patient messages demonstrate a markedly improved emotional state when contacting clinicians post-intervention as compared to pre-intervention. Additionally, both prescribing physicians and involved pharmacists consistently described decreases to clinician burden, improved experience & satisfaction, praise for their colleagues, and excellent coordination & efficiency.

While prior research has examined the “how” and “who” of PAs (how to utilize medical assistants, nurses, and pharmacists to help work through PAs after prescription submission), this study adds the “when” by involving pharmacists *before* prescription submission. This critical innovation, informed by a detailed understanding of prescriber workflows, is a novel approach that improves clinician efficiency and the experience of both patients and clinicians.

Further, the burden that PAs place on all types of clinical staff is well documented and quite concerning when viewed in the broader context of growing prescriber workloads and burnout. In addition to measuring the clinical impact of PAs on patients, it is imperative that clinician experience is studied in detail to better understand how interventions can be designed in ways that ease clinician burden. These results suggest that creative approaches to resource allocation can improve clinician experience as well as overall efficiency, presenting the case for a rare “win-win” that can positively impact both clinicians as well as patients.

Despite the robust multi-method analytic approach, study limitations include a relatively small sample size of each clinician group (eleven prescribers and seven pharmacists) from a single health center who participated in the qualitative evaluation. Similarly, patient messages were analyzed from a single health center and may not be generalizable to other populations or health systems. Additional studies with a wider range of clinicians or the potential use of clinician experience surveys would better reinforce these findings. Finally, as with all observational analyses, the potential for residual confounding remains and interpretation is limited only to association and not causation.

## Conclusions

Investing staff resources before, during, and after the prior authorization process greatly improves the clinician experience for both prescribers and pharmacists. It also increases the positive valence of patient portal messages, suggesting an improvement in patient experience. These findings should inspire health systems to seek other creative solutions to medication access and the PA process.

## Supporting information

Supplementary Material and eTables

## Data Availability

All data produced in the present work are contained in the manuscript and related supplemental materials.

## Acknowledgements

None

