## Supplementary Material and eTables for "Implementation of an Enhanced Medication Access Workflow within a Health System Specialty Pharmacy: Impact on Patient & Clinician Experience"

### Semi-structured Interview Methods

Interviews were conducted by two experienced researchers (TC, VG) between September 2022 and May 2023. Interviews were conducted through virtual video calls, lasted 30-60 minutes each, and were audio-recorded and transcribed verbatim.

Two qualitative researchers coded all transcripts (TC, VG) using inductive coding based on an adapted grounded theory approach.[i,ii] Coders completed independent selective, inductive coding of interview transcripts, using results to explore themes related to the intervention and its impact on clinician experience. Through the review and coding process, researchers iteratively developed and used a shared coding framework that identified emergent themes and subthemes. Data were organized, shared, and analyzed using MAXQDA analysis software and amended through discussion. This process, the results, and preliminary theory were shared with key stakeholders confirm that researchers' interpretations and related themes aligned with real world practice experience. This analytical plan supported our efforts to maximize the validity and trustworthiness of these data.

### Quantitative Evaluation Methods of Patient Messages

We scored the emotional valence of each of these messages by adapting TweetEval a machine learning algorithm originally trained on Twitter data to score portal messages sent by patients.[iii,iv,v] This model probabilistically classifies the emotional valence of each message as positive, neutral, or negative. This model was developed to score the emotional valence of 160-character tweets. We adapted it for patient portal messages by scoring each line of the message separately and averaging the results. As needed, lines longer than 160 characters were broken into sentences using spaCy and the line was scored using the average of the sentence level scores.[vi] We summarized the probability scores from this model using the *net positive valence* which we define as the probability that a message has a positive valence less the probability it has a negative valence. We scaled the net positive valence by 100 to aid interpretation and it therefore ranges from -100 (most negative) to 100 (most positive).

**eTable 1.** *Clinical and demographic characteristics of all patients first prescribed a specialty medication or referred to specialty pharmacy.*

| Characteristic | Overall, N (%) | Pre, N (%) | Post, N (%) | Difference, % (95% CI) | Adjusted[1] Difference, % (95% CI) | p-value |
|---|---|---|---|---|---|---|
| Total | 6,888 (100) | 2,945 (100) | 3,943 (100) | - | - | - |
| Age in years, mean (IQR) | 47.9 (35.3-59.5) | 48.5 (36.0-60.1) | 47.1 (34.4-58.8) | -1.4 (-2.2 to -0.7) | -0.7 (-1.4-0.7) | 0.08 |
| Female | 4,838 (70.2) | 2,729 (69.2) | 2,109 (71.6) | 2.4 (0.2-4.6) | 1.8 (-0.3-4.0) | 0.09 |
| *Self-reported Race* | | | | | | |
| American Indian and Alaska Native | 41 (0.6) | 27 (0.7) | 14 (0.5) | -0.2 (-0.6-0.1) | -0.2 (-0.5-0.2) | 0.33 |
| Asian | 158 (2.3) | 78 (2.0) | 80 (2.7) | 0.7 (0.0-1.5) | 0.9 (0.2-1.7) | 0.038 |
| Black | 574 (8.3) | 347 (8.8) | 227 (7.7) | -1.1 (-2.4-0.2) | -0.4 (-1.7-0.9) | 0.54 |
| White | 5,685 (82.5) | 3,269 (82.9) | 1,146 (82.0) | -0.9 (-2.7-0.9) | -2.1 (-3.9 to -0.3) | 0.026 |
| Other[3] | 342 (5.0) | 179 (4.5) | 163 (5.5) | 1.0 (-0.1-2.0) | 1.2 (0.1-2.3) | 0.029 |
| Unknown[4] | 88 (1.3) | 43 (1.1) | 45 (1.1) | 0.4 (-0.1-1.0) | 0.5 (-0.0-1.1) | 0.063 |
| *Ethnicity* | | | | | | |
| Hispanic | 228 (3.3) | 139 (3.5) | 89 (3.0) | -0.5 (-1.3-0.4) | -0.4 (-1.2-4.8) | 0.39 |
| Non-Hispanic | 6,495 (94.3) | 3,714 (94.2) | 2,781 (94.4) | 0.2 (-0.9-1.3) | 0.1 (-1.0-1.2) | 0.87 |
| Unknown[3] | 165 (2.4) | 90 (2.3) | 75 (2.5) | 0.3 (-0.5-1.0) | 2.8 (-4.7-1.0) | 0.73 |
| *Provider Specialty* | | | | | | |
| Dermatology | 908 (13.2) | 587 (14.9) | 321 (10.9) | -4.0 (-5.6 to -2.4) | - | - |
| Gastroenterology, Irritable Bowel Disease | 860 (12.5) | 392 (9.9) | 468 (15.9) | 5.9 (4.3-7.6) | - | - |
| Neurology[3] | 2,503 (36.3) | 1,348 (34.2) | 1,155 (39.2) | 5.0 (2.7-7.3) | - | - |
| Pulmonology | 520 (7.5) | 364 (9.2) | 156 (5.3) | -3.9 (-5.1 to -2.7) | - | - |
| Rheumatology | 2,097 (30.4) | 1,252 (31.8) | 845 (28.7) | -3.1 (-5.2 to -0.9) | - | - |

[1]Adjusted for provider specialty.
[2]Asian includes: "Asian", "Asian Indian", "Chinese", "Korean", "Filipino" and "Other Asian".
[3]Other includes: "Middle Eastern/North African", "Native Hawaiian", "Native Hawaiian and Other Pacific Islander", "Multi" and "Other". Excepting "Multi" and "Other" groups with fewer than 11 patients overall were included here to protect patient privacy.
[4]Unknown includes: "Choose not to disclose", "Patient Refused", "Unknown", and missing values (17 for ethnicity).
[5]Provider specialty is imbalanced due to stepped rollout of the specialty pharmacy referral.
[6]Neurology includes neurologists specializing in both headache and multiple sclerosis.
Abbreviations: CI = Confidence Interval, IQR = Interquartile Range

**eTable 2.** *Clinical and demographic characteristics of patients in the post period, stratified by use of specialty referral.*

| Characteristic | No Referral, N (%) | Referral, N (%) | Difference, % (95% CI) | Adjusted[1] Difference, % (95% CI) | p-value |
|---|---|---|---|---|---|
| Total | 2,415 (100) | 2,131 (100) | - | - | - |
| Age in years, mean (IQR) | 47.7 (35.2-59.6) | 46.5 (34.1-58.2) | -1.2 (-2.1 to -0.3) | -0.2 (-1.1 to 0.7) | 0.68 |
| Female | 1,707 (29.3) | 1,561 (570) | 2.6 (-0.0 to 4.2) | 1.7 (-0.1 to 4.3) | 0.19 |
| *Self-reported Race* | | | | | |
| American Indian and Alaska Native | 17 (0.7) | 9 (0.4) | -0.3 (-0.7 to 0.2) | -0.2 (-0.6 to 0.2) | 0.25 |
| Asian | 47 (1.9) | 56 (2.6) | 0.7 (-0.2 to 1.6) | 1.0 (0.5-1.9) | 0.022 |
| Black | 169 (7.0) | 134 (6.3) | -0.7 (-2.2 to 0.7) | -0.3 (-1.7 to 1.2) | 0.72 |
| White | 2,060 (85.3) | 1,791 (84.0) | -1.3 (-3.4 to 0.8) | -2.3 (-4.4 to -0.2) | 0.033 |
| Other[3] | 98 (4.1) | 115 (5.4) | 1.3 (0.1-2.6) | 1.5 (0.3-2.8) | 0.016 |
| Unknown[4] | 34 (1.0) | 26 (1.2) | 0.2 (-0.4 to 0.8) | 0.2 (-0.4 to 0.1) | 0.47 |
| *Ethnicity* | | | | | |
| Hispanic | 82 (3.4) | 59 (2.8) | -0.6 (-1.6 to 0.4) | -0.5 (-1.5 to 0.5) | 0.32 |
| Non-Hispanic | 2,275 (94.2) | 2,023 (94.9) | 0.7 (-0.6 to 2.0) | 0.7 (-0.1 to 2.0) | 0.33 |
| Unknown[3] | 58 (2.4) | 49 (2.3) | -0.1 (-0.1 to 0.8) | -0.2 (-1.0 to 0.7) | 0.74 |
| *Provider Specialty* | | | | | |
| Dermatology | 319 (13.2) | 180 (8.4) | -4.8 (-6.6 to -3.0) | - | - |
| Gastroenterology, Irritable Bowel Disease | 301 (12.5) | 390 (18.3) | 5.8 (3.7-7.9) | - | - |
| Neurology[3] | 759 (31.4) | 850 (39.9) | 8.5 (5.7-11.2) | - | - |
| Pulmonology, Allergy | 222 (9.2) | 113 (5.3) | -3.9 (-5.4 to -2.4) | - | - |
| Rheumatology | 814 (33.7) | 598 (28.1) | -5.6 (-8.3 to -3.0) | - | - |

**eTable 3.** *Net positive valence of patient portal messages.*
Net positive valence for message sent by patients within 180 days of first referral to specialty pharmacy or prescription of a specialty pharmacy drug included in this study. This table presents the unadjusted means and interquartile ranges across all messages.

| | **Net Positive Valence** | |
|---|---|---|
| | **Pre, Mean (IQR)** | **Post, Mean (IQR)** |
| Overall | 7.8 (0.0-25.0) | 12.2 (0.0-33.3) |
| *Provider Specialty* | | |
| Dermatology | 8.4 (0.0-25.8) | 18.2 (0.0-50.0) |
| Gastroenterology, Irritable Bowel Disease | 12.3 (0.0-26.5) | 13.1 (0-0-30.0) |
| Neurology | 4.2 (0.0-25.0) | 10.1 (0.0-33.3) |
| Pulmonology, Allergy | 12.2 (0.0-31.2) | 12.8 (0.0-33.3) |
| Rheumatology | 7.4 (0.0-25.0) | 13.2 (0.0-33.3) |

IQR = Interquartile range or the 25th and 75th percentiles of the distribution.

**eTable 4.** *Adjusted net positive valence of patient portal messages and change from pre to post by specialty.*

| | **Net Positive Valence** | | | | |
|---|---|---|---|---|---|
| | **Pre, Mean (95% CI)** | **Post, Mean (95% CI)** | **Difference (95% CI)** | **Trend Adjusted Difference (95% CI)** | **Trend Adjusted Difference, Excluding 2020 (95% CI)** |
| Overall[1] | 7.8 (6.8-8.9) | 12.2 (11.1-13.3) | 5.3 (3.8-6.8) | 5.1 (3.3-7.1) | 3.9 (1.9-5.8) |
| *Provider Specialty* | | | | | |
| Dermatology | 9.5 (6.6-12.4) | 20.5 (16.6-24.5) | 11.0 (6.1-15.9) | 9.4 (-5.6-24.3) | 7.4 (-1.7-16.5) |
| Gastroenterology, Irritable Bowel Disease | 12.3 (9.5-15.5) | 14.0 (11.6-16.5) | 1.7 (-2.0 to 5.4) | -[2] | 0.1 (-4.9-5.3) |
| Neurology | 6.9 (5.0-8.7) | 13.2 (11.5-14.9) | 6.3 (3.8-8.9) | -[2] | 5.3 (2.1-8.5) |
| Pulmonology | 14.7 (11.3-18.0) | 22.2 (17.4-26.9) | 7.5 (1.7-13.3) | -[2] | 4.6 (-1.7-10.9) |
| Rheumatology | 9.6 (7.9-11.4) | 13.8 (11.7-15.8) | 4.2 (1.5-6.8) | -[2] | 3.3 (0.3-6.3) |

Abbreviations: CI = Confidence Interval
[1]The overall pre and post means are average predictions and the overall difference the average marginal effect. The other means and differences are estimated using contrasts of model coefficients.
[2] Same as the unadjusted difference.

**eTable 5.** *Sensitivity analysis assessing for trends in net positive valence of patient portal messages by specialty.* For each specialty, "common trend" refers to secular trend constant across the pre and post periods while "different specialty trends" refers to a pre implementation trend that is allowed to change post implementation. For each specialty, we tested for trends only for the model with the best AIC and only when that AIC was lower (better) than the AIC of the primary model.

| Model | # of Parameters | AIC | Best for Specialty | Likelihood Ratio Test, p |
|---|---|---|---|---|
| Primary | 12 | 69,155.7 | - | - |
| Primary + Common Dermatology Trend | 13 | 69,147.7 | Yes | 0.004 |
| Primary + Different Dermatology Trends | 14 | 69,148.4 | No | - |
| Primary + Common Gastroenterology Trend | 13 | 69,157.0 | No | - |
| Primary + Different Gastroenterology Trends | 13 | 69,158.1 | No | - |
| Primary + Common Neurology Trend | 14 | 69,156.7 | No | - |
| Primary + Different Neurology Trends | 13 | 69,158.1 | No | - |
| Primary + Common Pulmonology Trend | 14 | 69,155.6 | Yes | 0.14 |
| Primary + Different Pulmonology Trends | 13 | 69,156.9 | No | - |
| Primary + Common Rheumatology Trend | 14 | 69,157.0 | No | - |
| Primary + Different Rheumatology Trends | 13 | 69,158.5 | No | - |

AIC = Akaike Information Criterion

**eTable 6.** *Estimated parameters for the primary model and two sensitivity analyses.*

| | Estimate (Standard Error) | | |
|---|---|---|---|
| **Model Parameter** | **Primary** | **Primary + Dermatology Trend** | **Primary + Dermatology Trend, Excluding 2020**[1] |
| Intercept[1] | 9.5 (1.5) | 0.5 (3.2) | 11.4 (6.4) |
| Post Intervention | 11.0 (2.5) | 1.8 (3.8) | 7.4 (4.7) |
| Gastroenterology | 2.8 (2.0) | 11.8 (3.5) | 2.5 (6.8) |
| Gastroenterology x Post | -9.3 (3.1) | -0.1 (4.3) | -7.3 (5.4) |
| Neurology | -2.7 (1.7) | 6.3 (3.3) | -3.6 (6.5) |
| Neurology x Post | -4.7 (2.8) | 4.5 (4.0) | -2.1 (4.9) |
| Pulmonology | 5.2 (2.2) | 14.2 (3.6) | 5.9 (6.7) |
| Pulmonology x Post | -3.5 (3.9) | 5.7 (4.8) | -2.8 (5.4) |
| Rheumatology | 0.1 (0.2) | 9.9 (3.1) | -1.0 (6.5) |
| Rheumatology x Post | -6.9 (2.9) | 2.3 (4.1) | -4.1 (4.9) |
| Dermatology x Time[2] | - | 21.9 (6.9) | 2.0 (12.0) |
| Patient Random Effect, standard deviation | 20.4 | 20.3 | 20.4 |
| Residual, standard deviation | 40.3 | 40.3 | 42.3 |

[1]n = 48,393 messages from 3,595 patients.
[2]Represents the adjusted mean for reference specialty, Dermatology, prior to the intervention.
[3]Time is scaled to range from 0 at study start to 1 at study end.

**eTable 7.** *Specialty specific changes for the percentage of messages classified as having a positive emotional valence.* Messages were classified as positive if the net positive valence was greater than 75.

| | % of Messages Classified as Positive | | | |
|---|---|---|---|---|
| | **Pre[1], Mean (95% CI)** | **Post[1], Mean (95% CI)** | **Difference[2] (95% CI)** | **Difference[2], Excluding 2020 (95% CI)** |
| Overall | 4.7 (4.3-5.1) | 11.6 (10.9-12.3) | 6.9 (6.1-7.7) | 5.9 (5.0-6.9) |
| *Provider Specialty* | | | | |
| Dermatology | 11.3 (8.4-14.1) | 9.1 (6.8-11.4) | -2.2 (-6.4-2.1) | -1.2 (-6.8-4.5) |
| Gastroenterology, Irritable Bowel Disease | 2.9 (2.2-3.6) | 9.5 (8.2-10.8) | 6.6 (5.1-8.0) | 5.5 (3.8-7.2) |
| Neurology | 3.2 (2.7-3.7) | 12.2 (11.1-13.4) | 9.0 (7.8-10.3) | 8.1 (6.8-9.4) |
| Pulmonology | 5.6 (4.3-6.8) | 14.9 (11.6-18.2) | 9.3 (5.8-12.9) | 7.6 (4.1-11.2) |
| Rheumatology | 5.5 (4.8-6.2) | 11.9 (10.6-13.2) | 6.4 (4.9-7.9) | 5.2 (3.7-6.8) |

[1]Average adjusted predictions.
[2]Adjusted for pre-existing trend in Dermatology.

**eTable 8.** *Target Specialty Drug List*

| Generic Specialty Drug Names | | |
|---|---|---|
| abatacept | abrocitinib | adalimumab |
| anakinra | apremilast | atogepant |
| baricitinib | belimumab | benralizumab |
| brodalumab | certolizumab | dalfampridine |
| deucravacitinib | dimethyl fumarate | diroximel fumarate |
| dupilumab | ereumab | etanercept |
| fingolimod | fremanezumab | galcanezumab |
| glatiramer | golimumab | guselkumab |
| icatibant | lasmiditan | mepolizumab |
| omalizumab | ozanimod | pirfenidone |
| rimegepant | risankizumab | ruxolitinib |
| sarilumab | secukinumab | siponimod |
| tapinarof | tezepulumab | tocilizumab |
| tofacitinib | ubrogepant | upadacitinib |
| ustekinumab | | |

### REFERENCES

[i] Charmaz K. Constructing Grounded Theory: A Practical Guide Through Qualitative Analysis. Thousand Oaks, CA: Sage Publications; 2006.

[ii] Bulawa P. Adapting grounded theory in qualitative research: reflections from personal experience. *Int Res Educ.* 2014;2(1):145-168.

[iii] Barbieri F, Camacho-Collados J, Anke LE, Neves L. TweetEval: Unified Benchmark and Comparative Evaluation for Tweet Classification. 2020. Findings of the Association for Computational Linguistics: EMNLP 2020, pages 1644–1650, Online. Association for Computational Linguistics.

[iv] Twitter-roBERTa-base for Sentiment Analysis. https://huggingface.co/cardiffnlp/twitter-roberta-base-sentiment (accessed February 2024)

[v] TweetEval. https://cardiffnlp.github.io/project/tweeteval/ (accessed February 2024).

[vi] sPacy. https://spacy.io/ (accessed February 2024).
